# SARS-CoV-2 Surveillance in US Wastewater: Leading Indicators and Data Variability Analysis in the 2023-2024 Season

**DOI:** 10.1101/2024.08.28.24312739

**Authors:** Hannes Schenk, Wolfgang Rauch, Alessandro Zulli, Alexandria B. Boehm

## Abstract

Wastewater-Based Epidemiology (WBE) has become a powerful tool for assessing disease occurrence in communities. This study investigates the coronavirus disease 2019 (COVID-19) epidemic in the United States during the 2023-2024 season using wastewater data from 189 wastewater treatment plants in 40 states and the District of Columbia. Severe acute respiratory syndrome coronavirus 2 (SARS-CoV-2) and pepper-mild mottle virus normalized SARS-CoV-2 concentration data were compared with COVID-19 hospitalization admission data at both national and state levels. We further investigate temporal features in wastewater viral abundance, with peak timing and cross-correlation lag analyses indicating that wastewater SARS-CoV-2 concentrations precede hospitalization admissions by 2 to 12 days. Lastly, we demonstrate that wastewater treatment plant size, assessed by number of population served, has a significant effect on the variability of measured SARS-CoV-2 concentrations. This study highlights the effectiveness of WBE as a non-invasive, timely and resource-efficient disease monitoring strategy, especially in the context of declining COVID-19 clinical reporting.

## 1. Introduction

On 30 January 2020, the World Health Organization (WHO) declared the coronavirus disease 2019 (COVID-19) outbreak a public health emergency of international concern [1]. Three years and three months later, the WHO declared the end of the public health emergency, despite Severe acute respiratory syndrome coronavirus (SARS-CoV-2) infections remaining a leading cause of death worldwide and in the United States (US). Even with the availability of vaccines and therapeutic treatments in the US, SARS-CoV-2 was responsible for a reported 49,931 deaths in 2023, highlighting the need to understand COVID-19 disease burden to inform public health policies [2]. While clinical data remain the standard for tracking disease burden, maintaining testing on a large scale is resource intensive, fails to detect asymptomatic cases and relies on the compliance of the public [3]. Wastewater-based epidemiology (WBE) provides a supplementary monitoring option that helps fill knowledge gaps such as undetected community spread, asymptomatic cases, and the lag in clinical reporting [4–6]. Human shedding of pathogens and chemicals into wastewater provides an important source of information on the health of the entire community living in a catchment area [7]. As an established surveillance method, WBE contributed to the management of the COVID-19 pandemic early on. Medema *et al*. [8], Ahmed *et al*. [9] among others ([10–14]) laid out the foundational work for SARS-CoV-2 surveillance using wastewater. While clinical testing efforts have decreased, WBE remains a central technology in monitoring SARS-CoV-2 in the population, as well as increasing in use for the detection of other pathogens [15–18].

Timely epidemiological data are crucial for assessing infectious disease outbreaks and implementing the necessary public health interventions. Both clinical testing data and hospital admission data correlate strongly to viral concentrations in wastewater, with wastewater leading both clinical testing and hospitalization data [14, 19–22]. WBE as an early warning system has been discussed in literature thoroughly [23, 24]. A streamlined process of logistics, sample analysis and data reporting are critical to leverage the temporal advantages of WBE. Understanding the lead times of WBE data is critical for the construction of forecasting models.

In this study the 2023-2024 COVID-19 epidemic in the US is investigated by analyzing longitudinal measurements of SARS-CoV-2 RNA in wastewater from 189 wastewater treatment plants (WWTPs) throughout the US. The data are aggregated on several spatial levels to compare to data on COVID-19 hospitalizations in 10 states. This paper’s novelty lies in its extensive dataset from WWTPs across the US, providing a comprehensive nationwide analysis of the 2023-2024 post-pandemic period. We then investigate leading or lagging behavior by comparing peak timings and computing maximum cross-correlation coefficients. Lastly, we demonstrate that SARS-CoV-2 RNA concentration variability is a function of WWTP size, offering new insights into the influence of WWTP size on the underlying dynamics.

## 2. Methods

### 2.1 Viral quantification and data characterization

For this study, wastewater data and hospitalization data are analyzed. COVID-19 hospitalization data are publicly available from the Centers for Disease Control and Prevention (CDC) [2]. State-aggregated, daily hospitalization data consists of data on COVID-19 occupancy and admission numbers. The wastewater data used in this study were retrieved through the nucleic acid extraction of settled solids from WWTPs nationwide. In June 2024 a total of 189 treatment plants were monitoring SARS-CoV-2 RNA in 40 different states throughout the US using a consistent approach by a single laboratory. Wastewater composite samples are collected with a sample frequency of 2 to 3 times per week for most plants, while some plants collect samples up to 7 times per week. Settled solids were extracted after dewatering by centrifugation at 24,000 x g for 30 minutes [25]. Solids were resuspended in DNA/RNA shield to a concentration of 75 mg/mL. Bovine coronavirus (BCoV) was used as a positive recovery control in all samples. Extraction was performed using a Chemagic Viral DNA/RNA 300 kit H96 in conjunction with a Perkin Elmer Chemagic 360 (Chemagic #CMG-1033-S). Inhibitor removal was performed using a Zymo OneStep-96 PCR Inhibitor removal kit (Zymo Research #D6035). Extraction negative controls and positive controls were extracted simultaneously. SARS-CoV-2 digital droplet RT-PCR was performed using primers and probes previously described [25]. BCoV and pepper-mild mottle virus were quantified in a duplex assay in each sample as controls. Each sample was run in 6 to 10 replicate wells and merged before analysis [26]. In the study period between 1^st^ of May 2023 and 1^st^ of June 2024, a total of 29,364 daily samples were examined. Readers are directed to a Data Descriptor for a full description of the SARS-CoV-2 measurements methods [27].

Figure 1 illustrates a map of the US, highlighting the number of WWTPs participating in SARS-CoV-2 RNA wastewater surveillance for the project. California, Texas, and Florida are the states with the highest number of contributing WWTPs, with 57, 14, and 13 plants respectively. Overall, 40 states have at least one WWTP monitoring SARS-CoV-2. Table 1 provides a detailed list of the states in the US, including the number of WWTPs contributing to the study. It also includes the total population served and the percentage of population coverage in each state.

**Table 1:** Number of WWTPs by state and percentage of population covered.

| State | #WWTPs | Pop.<br>served/10<sup>3</sup> | Pop.<br>coverage % | State | #WWTPs | Pop.<br>served/10<sup>3</sup> | Pop.<br>coverage % |
| --- | --- | --- | --- | --- | --- | --- | --- |
| CA | 57 | 20,511 | 52.6 | UT | 2 | 715 | 20.9 |
| TX | 14 | 2,425 | 7.9 | TN | 2 | 700 | 9.8 |
| FL | 13 | 3,650 | 16.1 | OH | 2 | 539 | 4.6 |
| GA | 8 | 1,109 | 10.1 | LA | 2 | 383 | 8.4 |
| NJ | 6 | 1,882 | 20.3 | NE | 2 | 300 | 15.2 |
| HI | 6 | 858 | 59.8 | IL | 2 | 149 | 32.7 |
| MI | 6 | 482 | 4.8 | MD | 2 | 145 | 1.2 |
| AL | 5 | 627 | 12.3 | NY | 2 | 120 | 2.3 |
| IN | 5 | 322 | 4.7 | CO | 2 | 60 | 0.6 |
| KS | 5 | 267 | 9.1 | MS | 2 | 53 | 1.0 |
| ME | 5 | 185 | 13.3 | NV | 1 | 990 | 1.8 |
| IA | 5 | 129 | 4.0 | KY | 1 | 423 | 31.0 |
| MN | 4 | 326 | 5.7 | AK | 1 | 220 | 9.4 |
| PA | 3 | 361 | 2.8 | CT | 1 | 140 | 3.9 |
| ID | 3 | 345 | 17.6 | WV | 1 | 100 | 5.6 |
| NC | 3 | 167 | 3.1 | WI | 1 | 44 | 0.7 |
| VA | 3 | 153 | 1.8 | SD | 1 | 20 | 2.2 |
| NH | 3 | 79 | 5.6 | AR | 1 | 15 | 0.5 |
| VT | 3 | 56 | 8.7 | DE | 1 | 13 | 1.3 |
| MA | 2 | 2,650 | 37.8 | WA | 1 | 10 | 0.1 |

**Figure 1:**
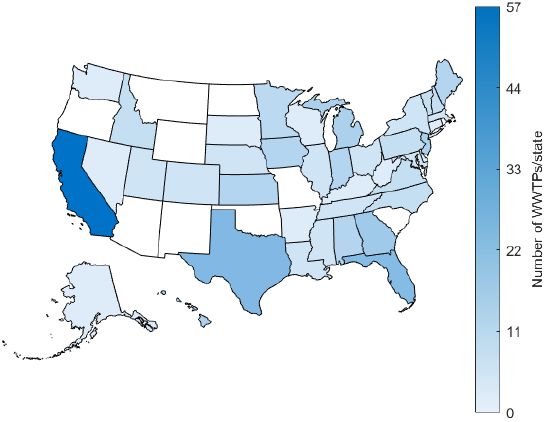
Map of the US. Shading indicates the number of WWTPs contributing to WBE in each state.

### 2.2. Data Pretreatment

SARS-CoV-2 (SC2) RNA and pepper-mild mottle virus (PMMoV) RNA concentrations were measured with digital droplet RT-PCR and reported as gene copies per gram dry weight. PMMoV is shed by humans in great abundance following the consumption of bell pepper and other pepper products [28]. Dividing SC2 by PMMoV concentrations compensates for the diversity of fecal strength of waste stream. This concept follows the mass balance model that relates concentrations of SARS-CoV-2 RNA in wastewater solids to incident infections of individuals in the sewershed [29]. The PMMoV normalized SC2 concentration is computed as follows:

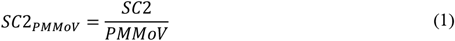

Before the spatial aggregation of WBE data, raw data were examined for outliers. Wastewater data are marked by random and systematic errors. Random errors are immanent to the technique of WBE and are caused with heterogeneities in the environmental sample and processes that affect concentrations in the sample; these can be difficult to reduce. Systematic errors are caused by a failure in the measurement process. With the outlier removal approach in this work, systematic errors are targeted. Concentrations larger than 3 standard deviations above the log_10_ transformed mean of the entire dataset (n=29,364) are discarded.

### 2.3. Spatial data aggregation

In order to compare wastewater data with hospitalization levels on a state by state basis (or on national scale), the SC2 concentration measurements were spatially aggregated. The spatial aggregation for WBE data in a state is performed by computing weighted daily averages of all WWTPs that provided data at a given date in that state, where the weighting factor is the population size that each plant serves. This computation results in a representative daily average of the SC2 circulation in the state, where the size of the plant is taken into consideration accordingly. The state aggregated daily weighted averages are calculated by

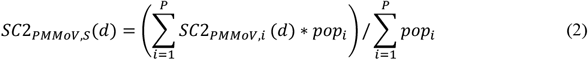

where the summation over the *P* indicates the plants in state S and *pop*_i_ denotes the population served by plant *i*. Analogous to the spatial aggregation on a state level, national weighted daily averages are computed by utilizing all available plants in the US. To obtain gapless time-series for the temporal analysis of the data, linear interpolation is performed if no datapoint is available at a given day after spatial aggregation.

### 2.4. Temporal analysis

The temporal features of SC2 normalized by PMMoV concentrations in wastewater are investigated in this study and compared on a state by state basis to hospitalization admission. The association between WBE data and hospitalization admission is determined using two approaches. First, cross-correlation function analysis (CCF) and second by examining the peak timing of the waves. Waves are periodic surges or peaks in the concentration of SC2_PMMoV_ over time. These waves represent fluctuations in SARS-CoV-2 RNA concentrations in wastewater. Furthermore, Spearman correlation *r* is examined to outline the qualitative relation between the time series.

Peak timing in time series provides a good reference point for comparison. The COVID-19 epidemic in the US in the season 2023-2024 is characterized by two local peaks. A smaller peak in fall 2023 and a more pronounced peak in December/January. In this work the peak timings for hospitalization admission and SC2_PMMoV_ in wastewater are compared relative to one another on a state by state basis. Peaks are determined by locating the highest values of the 7-day moving mean of the SC2_PMMoV_ concentrations in wastewater and hospitalization admission time-series. The peaks are determined for both occurring waves, where the 1^st^ of November is the date of separation between first and second wave. This date was chosen by visual inspection of the data and allows for a good separation of the two peaks for all states. The average time differential 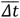 is than calculated by averaging the difference between peak occurrences of the two peaks for each state. 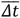 is calculated by

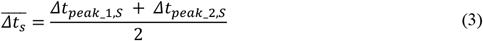

where 𝛥_*tpeak_1,S*_ and 𝛥_*tpeak_2,S*_ denote the time difference in days between the peaks of hospitalization admission and SC2_PMMoV_ concentrations for the two respective waves one and two. The subscript *S* denotes the state. The hospitalization peak date *t*_hosp_ is subtracted from the wastewater peak date t_ww_, so that negative days signify an earlier peak date in wastewater

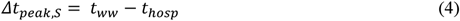

The emphasis in this analysis is on states with a population coverage of 15% or more (10 states). We assume that this constraint ensures the representativeness of wastewater data for SARS-CoV-2 circulation. All analyses are performed with MATLAB 2023b, The MathWorks Inc.

### 2.5. Data dispersion analysis

SC2 and PMMoV concentrations - and as a result therefrom the normalized SC2_PMMoV_ - in wastewater are characterized by substantial amounts of variability. Herein, data dispersion and variability characteristics are examined to quantify WBE data attributes. Data variability is explored for different sizes of WWTPs, where the proxy for plant size is given by the number of populations that each plant serves. The population served by the plants varies significantly, with the smallest plant serving approximately 5,000 individuals and the largest, a plant in Los Angeles, California, serving 4 million individuals. This analysis aims to investigate whether there are significant differences in data properties between small and large plants. It is hypothesized that differences in concentration variability may be observed due to the substantial variation in plant size, spanning three orders of magnitude.

To investigate the potential differences in concentration behavior between small and large wastewater treatment plants, the WBE dataset is partitioned into five groups. The partitioning regime is determined by quantile intervals of the population served. Data groups corresponding to the five quantile intervals are denoted as Q_0-0.2_, Q_0.2-0.4_, …, Q_0.8-1_ (from smallest 20% of plants to largest 20% of plants). Table 2 outlines the quantile intervals and data partitioning regime used for this analysis. The grouping in the described manner is designed to partition plants into groups that have similar sizes.

**Table 2:** Data partitioning into quantile ranges of plant sizes.

| Quantile ranges | Population served | #WWTPs | SC2 data points |
| --- | --- | --- | --- |
| $Q_{0-0.2}$ | 0-30,000 | 42 | 6278 |
| $Q_{0.2-0.4}$ | 30,001-64,000 | 34 | 4697 |
| $Q_{0.4-0.6}$ | 64,001-102,125 | 38 | 5632 |
| $Q_{0.6-0.8}$ | 102,126-227,238 | 38 | 6324 |
| $Q_{0.8-1}$ | 227,239-400,000 | 38 | 6433 |

For each of the five data groups standard deviation (SD) and interquartile-ranges (IQR) are computed of the log10 SC2 concentrations. This enables a comparison of the degree of variability as a function of plant size. To test that the five data groups stem from different statistical populations, two-sided Wilcoxon rank sum tests are performed between adjacent data groups.

## 3. Results

The COVID-19 epidemic in the timeframe May 2023 to June 2024 showed seasonal waves, similar to previous years [30]. This is shown in figure 2, plotting national aggregated daily SC2_PMMoV_ concentrations (and its 7-day moving average) along with hospitalization admissions in the US. This figure outlines the general development of the epidemic in the US in the studied timeframe. The two peaks are well pronounced in the national aggregated data. Reporting of SC2 hospitalization data is discontinued from the beginning of May 2024 and therefore truncated in time in figure 2. The bottom bar chart outlines the number of WWTPs that are monitored at a particular day. On the right of the figure, a scatter plot depicts WBE and hospitalization admission data with a simple ordinary least squares (OLS) regression line.

**Figure 2:**
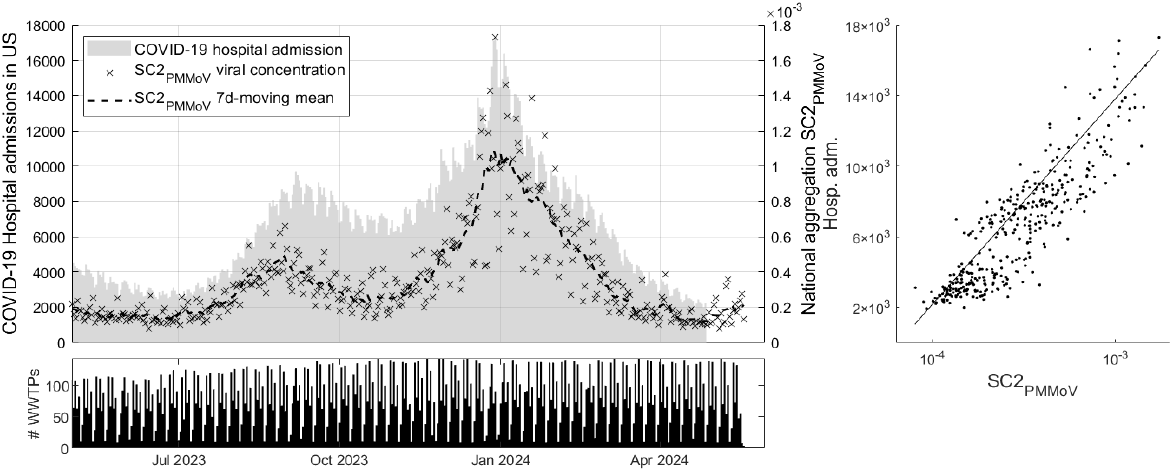
National aggregation of SC2_PMMoV_ and COVID-19 hospitalization admissions (top left), number of WWTPs measured per day (bottom) and OLS regression between the datasets.

Figure 3 displays the histogram of all SC2_PMMoV_ data in the timeframe May 2023 to June 2024. Values above 3 standard deviations above the mean are discarded as systematic errors (27 data points out of 29,364). The mean normalized concentration is 0.00052 and the outlier threshold is 0.012. Three standard deviations above the mean on the log_10_ transformed data corresponds to a 24-fold higher concentration on linear scale in relation to the mean of the data.

**Figure 3:**
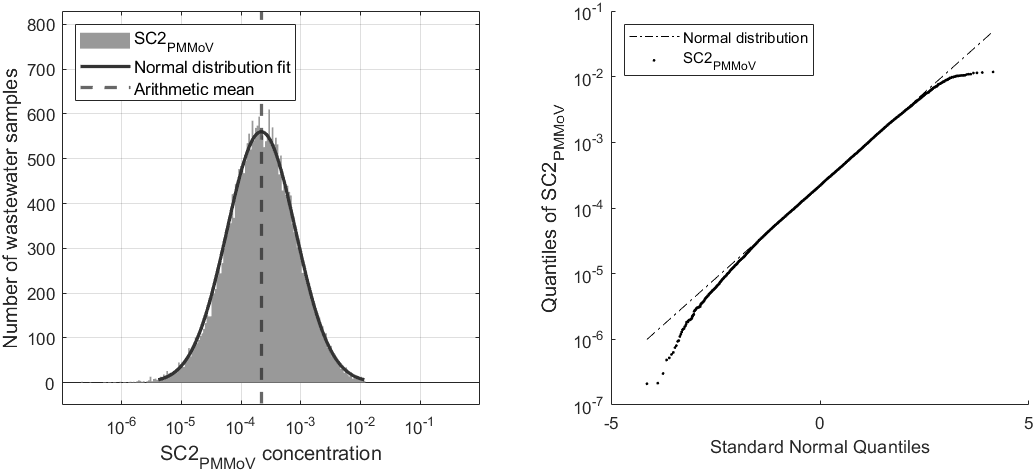
SC2_PMMoV_ data distribution and QQ-plot.

In addition to the histogram on figure 3, a normal distribution fit is computed to outline the resemblance of the SC2_PMMoV_ data with a log-normal distribution. To test the log_10_ transformed data for normality, Shapiro-Wilk and Kolmogorov-Smirnov tests are performed with a 5% significance level each. Both tests reject the null hypothesis and suggest that the data are not normally distributed. Compared to an ideal log-normal distribution, the measured data are characterized by a fat tail on the left. Unlike the right side of the distribution, the left side is not truncated with a lower bound for outlier removal. Low/very low concentration values are not considered outliers. On the right graph of figure 3, the quantile-quantile (QQ) plot is depicted. It can be seen that both tails of the distribution deviate from the normal line. The data are characterized by a slight negative skew (skewness=-0.16).

### 3.1. Temporal analysis results

In epidemiological surveillance, early detection and rapid information processing are critical. Temporal features of WBE SC2 monitoring, such as peak timing, cross-correlation lag and general wave development are analyzed. While other diseases like influenza or respiratory syncytial virus are characterized by clear onset/offset dates, SC2 is consistently circulating in the population since its outbreak in 2020 [31]. Therefore, peak occurrence timing is considered as the temporal feature for comparison. The COVID-19 epidemic in the US in the timeframe 2023-2024 was characterized by two waves. The peak of the first wave was less pronounced in SC2 concentration and hospitalization magnitude and occurred for most states in September 2023. The second wave occurred around January 1^st^ 2024. Figure 4 shows the cumulative occurrence of peaks in wastewater and hospitalization data for each state and for both waves.

**Figure 4:**
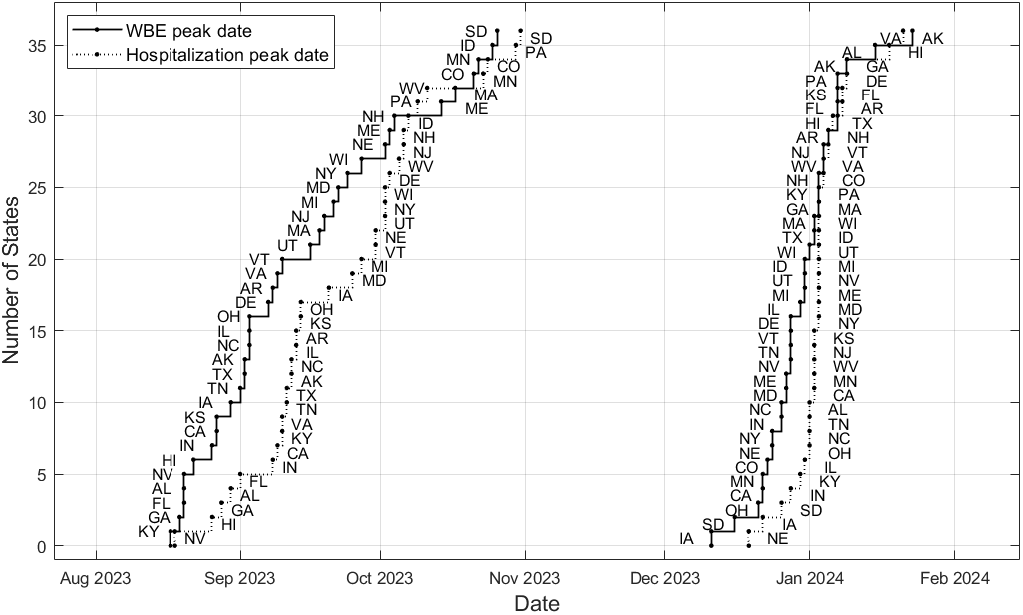
Cumulative number of States by peak occurrence, WBE and hospitalization admission. The abbreviation for each state is provided next to its data point.

It can be seen in figure 4 that the WBE peak generally occurs earlier than the hospitalization peak. The peak timings between hospitalization admission and SC2 concentrations visually decrease between the first and second wave. The second wave peaks across the US over a shorter time period. Reasons for this could be an increased infectivity of the COVID-19 variant and/or a rise in transmission due to more population mixing during the holiday travel. Spearman rank correlation of the order in which the states occur is 0.91 for the first wave and 0.56 for the second wave. This means that generally the order of occurrence of peaks is respected between the two data sets, especially in the first wave. The first wave occurs earliest in the Southeastern region including the states Kentucky, Georgia, Florida and Alabama, followed by Nevada, Hawaii and California among others. The second wave peaks earliest in midwestern states including Iowa, South Dakota, Minnesota, accompanied by Oklahoma, California and Nevada.

Figure 5 displays the state aggregated SC2_PMMoV_ concentrations and superimposed hospitalization admission per 100k population (gray bars) for states with a population coverage of 15% or more. SC2_PMMoV_ is shown on the left axis on log scale and hospitalization is shown on the right axis on linear scale.

**Figure 5:**
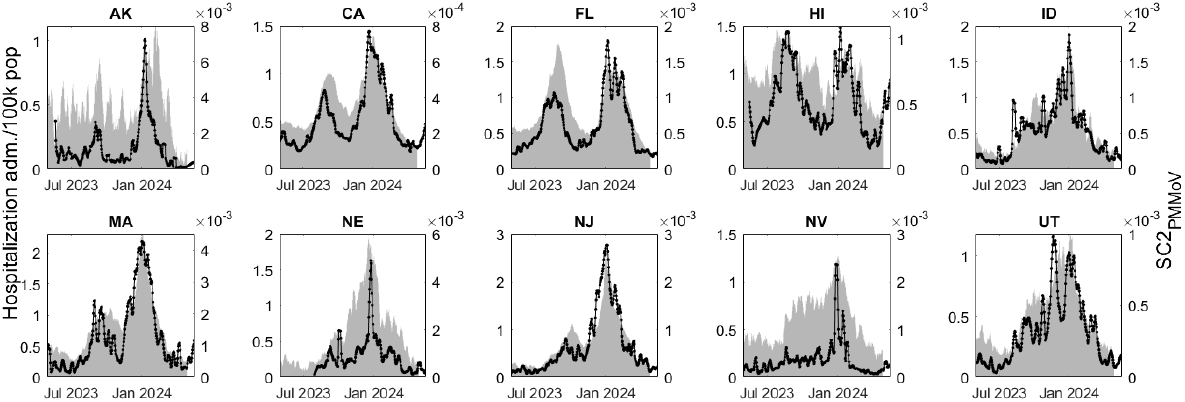
State aggregated SC2_PMMoV_ (black line) and hospitalization admission (gray bars).

Table 3 outlines the results of the temporal analysis. CCF lag between the time series (state aggregated SC2_PMMoV_ and hospitalization admission) and the time differential 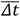 of the relative peak occurrence are listed. Negative values of CCF lag and 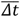 indicate that the WBE peak occurred before the hospitalization peak. Furthermore, Spearman correlation *r* values are listed as a comparative analysis between SC2 hospitalization admission and wastewater data for states with population coverage above 15%.

**Table 3:** SC2_PMMoV_ and hospitalization temporal quantitative feature comparison by state. Negative lag values indicate a time lead in wastewater over hospitalizations.

| State | $\overline{\Delta t}$ (d) | CCF lag | Spearman $r$ |
| --- | --- | --- | --- |
| CA | -12 | -3 | 0.88 |
| FL | -6.5 | -5 | 0.86 |
| HI | -7.5 | -10 | 0.62 |
| NJ | -7.5 | -4 | 0.93 |
| ID | 7.5 | -3 | 0.86 |
| MA | -12 | -4 | 0.83 |
| NE | 3.5 | 4 | 0.85 |
| UT | -9.5 | -5 | 0.86 |
| AK | -11 | -9 | 0.67 |
| NV | -2 | -2 | 0.76 |

SC2 in wastewater leads hospitalization admission in 8 out of 10 states, following the results of peak timing 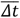. For the CCF lag, 9 out of 10 states show this characteristic. A median time lead of 4 and 7.5 days is observed for CCF lag and 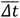 respectively among states with 15% or more population coverage. The correlation metrics *r* and *R*^2^ suggest a close agreement between hospitalization admission and SC2_PMMoV_ in wastewater (median *r*=0.85).

### 3.2. Data dispersion results

Differences in data dispersion characteristics for different plant sizes are observed. To examine the influence of plant size, the wastewater data are partitioned into 5 groups. The partitioning is governed by the quantiles of the population served by each plant and carried out as described in section 2.5.

Data dispersion results are listed in table 4 and visualized in figure 6. Table 4 describes data mean, median, standard deviation (SD) and interquartile range (IQR) of the partitioned data. The main measures of variability, SD and IQR, are observed to decrease with increasing plant size. This observation is in line with expectations, considering the more stochastic behavior of small plants and the law of large numbers. An intuitive explanation can be provided by considering a case prevalence of 0.1%. In a small plant serving 10,000 people, 10 individuals would be infected. Due to the size of the sewer system and the stochastic shedding behavior of these 10 infected individuals, SC2 concentrations may exhibit significant variability. Conversely, in a large WWTP serving a population of 1 million, 0.1% prevalence would correspond to 1,000 infected individuals. With a significant number of individuals shedding the virus, a more consistent discharge of the virus into the sewer system is likely. These findings align with the results from Nauta *et al*. [32], who performed Monte-Carlo simulations to estimate SC2 concentrations and data variability.

**Table 4:** Data dispersion properties log_10_(SC2) mean, median, standard deviation and interquartile range by plant size.

| Quantile | mean<br>$\log_{10}(\text{SC2})$ | median<br>$\log_{10}(\text{SC2})$ | SD<br>$\log_{10}(\text{SC2})$ | IQR<br>$\log_{10}(\text{SC2})$ |
| --- | --- | --- | --- | --- |
| $Q_{0-0.2}$ | 4.846 | 4.862 | 0.569 | 0.781 |
| $Q_{0.2-0.4}$ | 4.875 | 4.877 | 0.538 | 0.735 |
| $Q_{0.4-0.6}$ | 4.858 | 4.854 | 0.504 | 0.687 |
| $Q_{0.6-0.8}$ | 4.947 | 4.942 | 0.477 | 0.643 |
| $Q_{0.8-1}$ | 5.001 | 5.026 | 0.457 | 0.612 |

**Figure 6:**
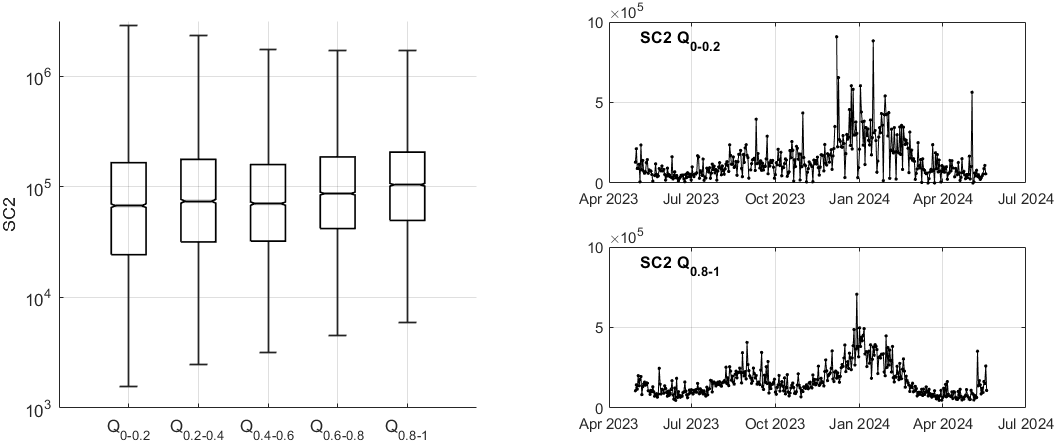
Data variability by quantile grouped data of different WWTPs sizes.

Figure 6 visualizes the data dispersion as a function of plant size. On the left, a boxplot diagram displays data median, upper and lower quartiles and minimum/maximum values by whiskers. The graph shows consistent decrease of data range and variability with the increase in plant size. On the right of figure 6 aggregated time-series are graphed, corresponding to Q_0-0.2_ (top, small plants) and Q_0.8-1_ (bottom, big plants). The ordinate axes are scaled equally for comparison.

To test the hypothesis that the partitioned wastewater groups based on plant size originate from statistically different data populations, two-sided Wilcoxon rank sum tests are performed. Four tests are carried out among the five groups between the adjacent groups. All tests reject the null hypothesis (that they stem from the same data population). All tests recommended to accept the alternative hypothesis (that they stem from different data populations) supports the thesis that there are underlying differences in data variability as a function of plant size. These findings can help public health officials interpret the significance of changes in WBE signals by taking into consideration the underlying population size contributing to the sewershed.

## 4. Conclusion

The COVID-19 epidemic in the US showed seasonality characteristics, that is periods of high and periods of low viral abundance in the population. The epidemic burden on the general population was lower – considering that case fatality was 60% lower in the studied timeframe, compared to the same season one-year earlier [2]. Since the initial outbreak in 2020, an average of two peak seasons per year have occurred [2]. The waves are observable in case data, hospitalization data and the SARS-CoV-2 RNA concentration in wastewater [30]. With the exception of the peak in January 2022 (which is the highest in the US), the weekly hospital admissions show decreasing peak amplitude over time for each consecutive occurring wave [2]. The increasing population contamination, the rise in vaccination levels and the changing SARS-CoV-2 variants with less severe symptoms led to greater resilience of society to the impacts of the epidemic.

This work investigates the SARS-CoV-2 RNA concentration data in US wastewater in the 2023-2024 season. Periodical waves characterizing the epidemic are observable. Clinical COVID-19 case reporting has largely been discontinued as of March 2024 [2, 33, 34]; hospitalization data reporting has been discontinued in early May 2024. In contrast, SARS-CoV-2 wastewater surveillance endeavors (among other pathogens) are well established and provide valuable information.

The work at hand examines statistical attributes of SARS-CoV-2 concentrations derived from wastewater surveillance. Firstly, temporal features, such as peak timing and CCF lag in the data are analyzed and compared to hospitalization admissions. The observations show that viral abundance in wastewater leads hospitalization admission between 2 and 12 days, in states with a population coverage of 15% or more. Data variability is analyzed and the influence of plant size on data dispersion has been observed, with the results demonstrating that smaller plants are subject to significantly more data variability. By partitioning the data into five batches based on plant size, a decrease in data variability with increased plant size is observed.

## Data Availability

All data produced are available online at https://purl.stanford.edu/hj801ns5929

## Acknowledgement

We acknowledge all the wastewater treatment plant staff who provided samples for this project.

## Notes

### Competing Interest Statement

The authors have declared no competing interest.

### Funding Statement

The study was partially supported by the Sergey Brin Family Foundation.

### Author Declarations

The study used only openly available human data that were originally located at https://covid.cdc.gov/covid-data-tracker/#datatracker-home

